# Core warming of coronavirus disease 2019 (COVID-19) patients undergoing mechanical ventilation – a protocol for a randomized controlled pilot study

**DOI:** 10.1101/2020.04.03.20052001

**Authors:** Nathaniel Bonfanti, Emily Gundert, Kristina Goff, Anne M. Drewry, Roger Bedimo, Erik Kulstad

## Abstract

**Background:** Coronavirus disease 2019 (COVID-19), caused by the virus SARS-CoV-2, is spreading rapidly across the globe, with no proven effective therapy. Fever is seen in most cases of COVID-19, at least at the initial stages of illness. Although fever is typically treated (with antipyretics or directly with ice or other mechanical means), increasing data suggest that fever is a protective adaptive response that facilitates recovery from infectious illness.

**Objective:** To describe a randomized controlled pilot study of core warming patients with COVID-19 undergoing mechanical ventilation.

**Methods:** This prospective single-site randomized controlled pilot study will enroll 20 patients undergoing mechanical ventilation for respiratory failure due to COVID-19. Patients will be randomized 1:1 to standard-of-care or to receive core warming via an esophageal heat exchanger commonly utilized in critical care and surgical patients. The primary outcome is the severity of acute respiratory distress syndrome (as measured by PaO2/FiO2 ratio) at 0, 24, 48, and 72 hours after initiation of treatment. Secondary outcomes include hospital and intensive care unit length of stay, duration of mechanical ventilation, viral load, and 30-day mortality.

**Results:** Resulting data will provide effect size estimates to guide a definitive multi-center randomized clinical trial. ClinicalTrials.gov registration number: NCT04426344.

**Conclusions:** With growing data to support clinical benefits of elevated temperature in infectious illness, this study will provide data to guide further understanding of the role of active temperature management in COVID-19 treatment and provide effect size estimates to power larger studies.

## Introduction

Traditionally, fever has been treated because its metabolic costs were felt to outweigh its potential physiologic benefit in an already stressed host.[1] However, increasing data suggest that fever may be a protective adaptive response that should be allowed to run its course under most circumstances.[2, 3] Higher early fever is associated with a lower risk of death among patients with an ICU admission diagnosis of infection.[4, 5] Fever may enhance immune-cell function,[6, 7] inhibit pathogen growth,[8-10] and increase the activity of antimicrobial drugs.[11] Fever potentially benefits infected patients via multiple mechanisms; in vitro and animal studies have shown that elevated temperatures augment immune function, increase production of protective heat shock proteins, directly inhibit microorganism growth, reduce viral replication, and enhance antibiotic effectiveness.[3, 12] More rapid recoveries are observed from chickenpox,[13] malaria,[14] and rhinovirus [15] infections with avoidance of antipyretic medication, and many innate and adaptive immunological processes are accelerated by fever.[16-18]

Randomized controlled trials have consistently failed to find benefits to treating fever of infectious etiology.[16, 19-24] A retrospective cohort study evaluating 1,264 patients requiring mechanical ventilation found that high fever (≥39.5°C) was associated with increased risk for mortality in mechanically ventilated patients; however, in patients with sepsis, moderate fever (38.3°C-39.4°C) was protective, and antipyretic medication was not associated with changes in outcome.[25] As recently as the 1910’s, the "malaria fever cure” (inducing fever to treat a range of conditions, an approach known as “pyrotherapy”) was widespread, with the originator of the idea receiving the Nobel Prize in Medicine or Physiology in 1927.[26, 27] Currently, the UK National Institute for Health and Care Excellence (NICE) recommend not using antipyretic agents “with the sole aim of reducing body temperature in children with fever.”[16, 28] Actively inducing hyperthermia by directly heating the body has been used in cancer treatment, with minimal adverse effects.[29-32] Hyperthermia has been found to have positive impacts on the immune system, causing increased levels of heat-shock proteins,[33, 34] which are directly related to antigen presentation and crosspresentation, activation of macrophages and lymphocytes, and activation and maturation of dendritic cells.[35] A pilot study of external warming of septic patients (ClinicalTrials.gov Identifier: <u>NCT02706275</u>) has recently been completed.

Many viruses replicate more robustly at cooler temperatures, such as those found in the nasal cavity (33-35°C) than at warmer core body temperature (37°C).[36-40] Coronavirus disease 2019 (COVID-19) and its causative virus (SARS-CoV-2) may behave similarly.[41] Simulations of the receptor binding domain (RBD) of SARS-CoV-2 found high flexibility near the binding site, suggesting that the RBD will have a high entropy penalty upon binding angiotensin-converting enzyme II (ACE2), and that consequently, the virus may be more temperature-sensitive in terms of human infection than other coronaviruses.[42] Notably, fever has often abated by the time a COVID-19 patient requires mechanical ventilation.[43] The aim of this study is to determine the effect of active core warming patients diagnosed with COVID-19 and undergoing mechanical ventilation. We hypothesize that active core warming will reduce the severity of acute respiratory distress syndrome, reduce the duration of mechanical ventilation, and improve survival compared to standard of care.

### Study Objectives

The purpose of the proposed pilot study is to determine if core warming improves respiratory physiology of mechanically ventilated patients with COVID-19, allowing earlier weaning from ventilation, and greater overall survival.

### Primary Objective

1. Measure the impact of esophageal core warming on severity of acute respiratory distress syndrome as measured by PaO2/FiO2 ratio 24 hours after initiation, and compare this to standard care.

### Secondary Objectives

1. Compare the duration of mechanical ventilation of patients treated with core warming to patients treated with standard care.
2. Compare the length of ICU and hospital stay of patients treated with core warming to patients treated with standard care.
3. Compare the viral shedding at day 7 and 14 of hospitalization between patients treated with core warming and patients treated with standard care.
4. Compare the 30-day mortality of patients treated with esophageal core warming to patients treated with standard care.

## Methods

This is a single-center pilot study to evaluate if core warming improves respiratory physiology of mechanically ventilated patients with COVID-19, allowing earlier weaning from ventilation, and greater overall survival. This prospective, randomized study will include 20 patients diagnosed with COVID-19, and undergoing mechanical ventilation for the treatment of respiratory failure. Patients will be randomized in a 1:1 fashion with 10 patients (Group A) randomized to undergo core warming with an esophageal heat transfer device, and the other 10 patients (Group B) serving as the control group. Patients randomized to Group A will have the esophageal heat transfer device placed in the ICU or other clinical environment in which they are being treated after enrollment and provision of informed consent from appropriate surrogate or legally authorized representative.

## Screening

Subjects will be recruited from the ICU or other clinical environment in which they are being treated (Emergency Department, step-down unit, etc.). Patients will be identified by the PI or other study investigators/coordinators as available. All patients without a DNR order with a diagnosis of COVID-19 and meeting inclusion criteria will be eligible for screening for any exclusion criteria. Written informed consent for the research study will be obtained from patient’s surrogate or legally authorized representative prior to enrollment.

## Study Intervention and Monitoring

Participants who have a signed research study consent form (via surrogate or legally authorized representative) will be randomized in a 1:1 fashion to core warming or to standard of care (standard temperature management and treatment). The esophageal heat transfer device will be used according to FDA 510(k) labeling (for patient warming). Patient temperature measurements will be collected for both the device and standard-of-care arms during the study period (up to 3 days). Device placement will be performed using standard protocol per instructions for use. The esophageal heat transfer device will be set to 42°C temperature after initial placement, and maintained at 42°C for the duration of treatment. All patients will have usual standard of care labs, vital signs, and imaging for patients in critical condition undergoing mechanical ventilation in the ICU. Specific parameters to be measured include PaO2 at regular intervals appropriate for patients undergoing mechanical ventilation, and FiO2 at the time of obtaining blood gases for PaO2 measurement, to allow calculation of P/F ratio.

## Study Endpoints

The purpose of this pilot study is to determine if core warming reduces the severity of acute respiratory distress syndrome as measured by PaO2/FiO2 ratio 0, 24, 48, and 72 hours after initiation.

## Primary Study Endpoints

*The primary endpoint of this study will be:*

1. PaO2/FiO2 ratio

*Secondary study endpoints include:*

1. Duration of mechanical ventilation
2. Duration of ICU and hospital stay
3. Amount of viral shedding
4. Patient mortality

## Inclusion Criteria

1. Patients above the age of 18 years old.
2. Patients with a diagnosis of COVID-19 on mechanical ventilation.
3. Patient maximum baseline temperature (within previous 12 hours) < 38.3°C.
4. Patients must have a surrogate or legally authorized representative able to understand and critically review the informed consent form.

## Exclusion Criteria

1. Patients without surrogate or legally authorized representative able to provide informed consent.
2. Patients with contraindication to core warming using an esophageal core warming device.
3. Patients known to be pregnant.
4. Patients with <40 kg of body mass.
5. Patients with DNR status.
6. Patients with acute stroke, post-cardiac arrest, or multiple sclerosis.

## Subject Recruitment

Subjects will be recruited from the ICU or other clinical environment in which they are being treated (Emergency Department, step-down unit, etc.). Patients will be identified by the PI or other study investigators/coordinators as available. All patients without a DNR order with a diagnosis of COVID-19 and meeting inclusion criteria will be eligible for screening for any exclusion criteria. Written informed consent for the research study will be obtained from patient’s surrogate or legally authorized representative prior to enrollment. If a patient enrolled in the study gains the capacity to consent for him/herself while the study is in progress, the patient will be approached by a study team member and the consent document will be presented directly to the patient. All questions the patient might have will be answered. The patient will be given the opportunity to either withdraw from the study or sign the consent form. The patient will be informed that his or her decision to withdraw from the study will not affect his or her medical care

## Duration of Study Participation

Participants will be involved for approximately 1 month, including screening, treatment, and follow-up. After consent, patient participation in the intervention phase will last up to 3 days for active treatment. The follow up for determination of outcome and duration of mechanical ventilation will occur at 1-month post-treatment. Additional data will be collected via chart review.

## Total Number of Subjects and Sites

This single-site study aims to recruit and randomize 20 patients. It is expected that up to 30 subjects may be consented in order to produce 20 randomized & evaluable subjects.

## Core temperature modulation

Core temperature control and warming will be performed with a commercially available esophageal heat exchange device (ensoETM, Attune Medical, Chicago, IL). This device is currently used world-wide for various patient temperature management goals, including post-cardiac arrest therapeutic hypothermia, [44-47] warming of burn patients,[48] warming general surgical patients,[49] cooling traumatic brain injury,[50] cooling heat stroke,[51] and the treatment of central fever.[52, 53] The device is a multi-chambered silicone tube placed in the esophagus and connected to a heat exchanger to provide heat transfer to or from a patient (video available at https://vimeo.com/306506411). Modulation and control of the patient’s temperature is achieved by adjusting setpoint on the external heat exchanger, which in turn controls the circulating water temperature. Two lumens of the device connect to the external heat exchanger, while a third central lumen provides stomach access for gastric decompression or tube feeding. It is a single-use, disposable, non-implantable device with an intended duration of use of 72 hours or less.

## Intervention Regimen

Patients who are randomized to core warming will have the esophageal heat transfer device placed in the ICU or other treatment area where patient is undergoing mechanical ventilation. The device will remain in place until the study is completed (24-72 hours). The device will be set to 42°C for the duration of the study period. It is expected that patient temperature will increase from baseline by 1°C to 2°C, but due to ongoing heat loss from the patient, the expected maximum patient temperature is below 39°C. The time course of illness of COVID-19 is such that most patients no longer have fever by the time of mechanical ventilation.[41] If patient temperature increases above this range and reaches 40°C, the device will be set to an operating temperature of 40°C, thereby preventing any further increase in patient temperature. Patient temperature will be followed at intervals per standard of care in the intensive-care setting for mechanically ventilated patients (typically hourly).

## Blinding

Due to the nature of this study, the physicians will not be blinded to the randomization assignment, however participants will be blinded. Once a subject is randomized, the research team will receive the randomization assignment (core warming or standard of care) and proceed with the procedures per the assignment.

### Data collection

- Demographics (including sex/gender, race, ethnicity, and age via date of birth)
- Past medical history, social history, physical exam findings and physicians notes
- Concurrent medications
- Physical exam
- Vital signs: temperature, blood pressure, heart rate, respiration rate, height and weight
- Clinical labs: complete blood count (CBC), chemistry profiles, liver function tests, inflammatory markers (CRP, ferritin), d-dimer, arterial blood gas for determination of PaO2
- Severity of illness: APACHE III, sequential organ failure assessment (SOFA) scoring systems
- Ventilator settings
- Pregnancy test for women of childbearing age
- Adverse events or unanticipated problems

Data will be collected via chart review, and is expected to be available from routinely obtained laboratory and vital sign data recorded at routine intervals (i.e., when labs are drawn for routine care in the ICU).

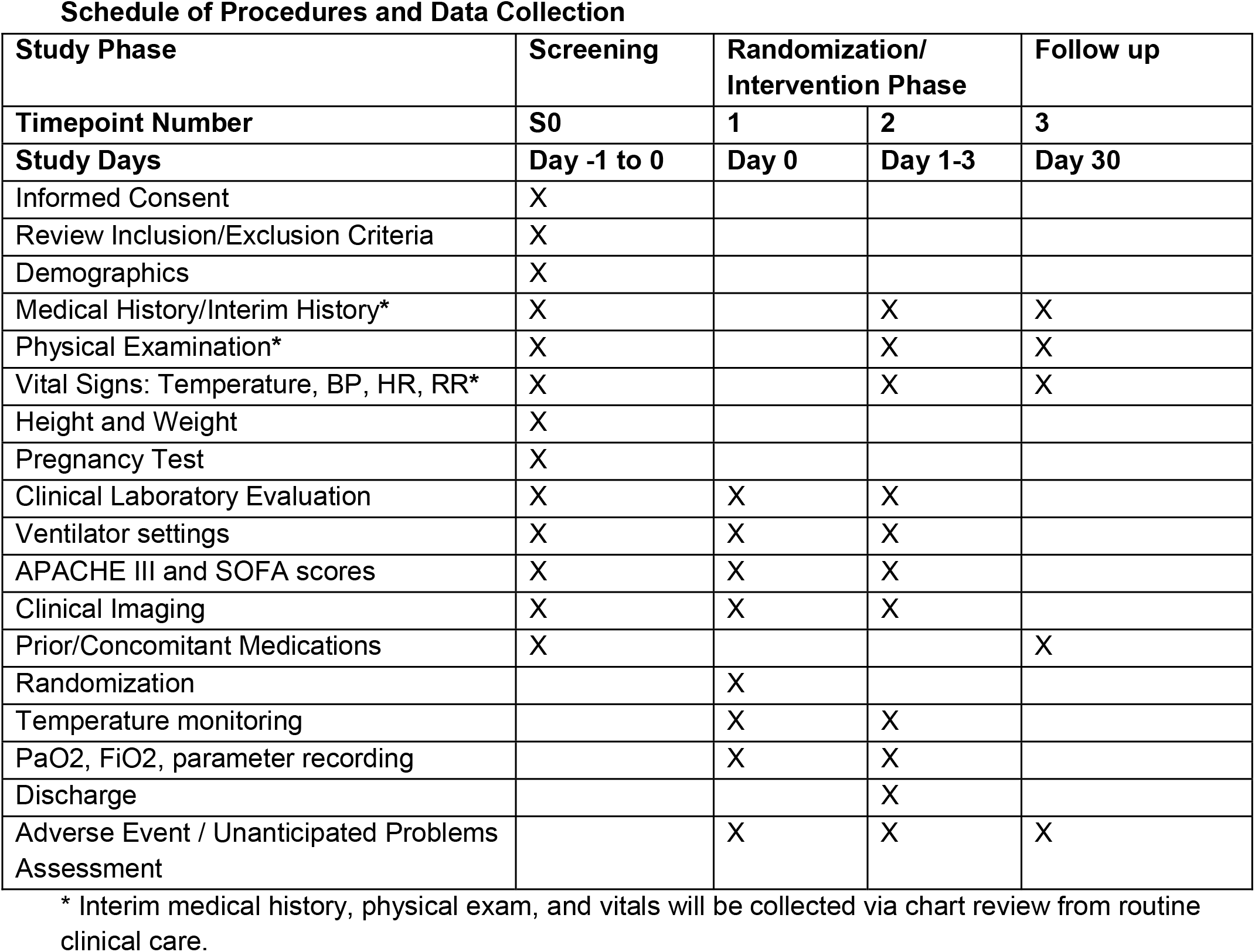
Schedule of Procedures and Data Collection.

### Statistical Plan

#### Primary Endpoint

The primary endpoint of this study will be the change in PaO2/FiO2 ratio 0, 24, 48, and 72 hours after implementation of core warming of ventilated patients. This endpoint will be compared between patients receiving core warming and those randomized to undergo standard care (standard temperature management, typically with antipyretics as needed).

#### Secondary Endpoints

Secondary endpoints include:

1. Duration of mechanical ventilation
2. Duration of ICU and hospital stay
3. Amount of viral shedding
4. Patient mortality

## Sample Size and Power Determination

Based on a prior study in patients with sepsis, a maximum temperature of 38.3°C to 39.4°C was associated with survival (aHR 0.61 [95% CI, 0.39-0.99]).[29] However, the effect of warming specific to COVID-19 patients remains uncertain, and as such, we are unable to accurately perform a power calculation for this pilot study. We believe that a total of 10 patients for each group will be required to yield the necessary pilot data to make an appropriate conclusion regarding the potential utility of core warming in improving pulmonary physiology, reducing mechanical ventilation duration, and increasing patient survival. It is anticipated that data from this pilot study can be used for planning future larger studies.

## Statistical Methods

We will utilize standard measures to report outcomes and measure differences between groups. Specifically, we will use descriptive statistics, including mean (standard deviation) and median (interquartile range). Normality will be assessed using histograms and the Kolmogorov–Smirnov test. Categorical variables will be compared using the chi-squared test or Fisher exact test. Continuous variables will be compared using the independent samples t or Mann–Whitney U test.

## Efficacy Analysis

This is a pilot study to determine the potential role of core warming during COVID-19 treatment.

## Interim Safety Analysis

All subjects entered into the study and randomized at the baseline timepoint will have detailed information collected on adverse events for the overall study safety analysis. An interim safety analysis will be performed after the first 10 subjects are enrolled in the trial. At this time the safety and tolerability of the study device will be assessed and if deemed safe and appropriate, enrollment will continue to 20 subjects.

## Subject Population for Analysis

All patients enrolled, randomized to a study arm, and completed in the study will be included for analysis.

## Conclusion

We describe, before the initiation of any data collection, our approach to obtaining and analyzing data from a pilot randomized-controlled trial of core warming patients undergoing mechanical ventilation due to COVID-19. We anticipate this framework will enhance the utility of the reported results and provide a solid basis from which to design and execute subsequent investigations.

## Data Availability

N/A

## Author contributions

NB, AD, RB, EK: conceptualization, design, drafting, and review of protocol and manuscript; EG, KG: protocol refinement, manuscript review and critical revision.

COI: EK declares equity interest in Attune Medical.

## References

1. Mohr NM, Doerschug KC: Point: Should antipyretic therapy be given routinely to febrile patients in septic shock? Yes. Chest 2013, 144(4):1096–1098.

2. Ray JJ, Schulman CI: Fever: suppress or let it ride? Journal of thoracic disease 2015, 7(12):E633–E636.

3. Drewry AM, Hotchkiss RS: Counterpoint: Should antipyretic therapy be given routinely to febrile patients in septic shock? No. Chest 2013, 144(4):1098–1101.

4. Saxena M, Young P, Pilcher D, Bailey M, Harrison D, Bellomo R, Finfer S, Beasley R, Hyam J, Menon D et al: Early temperature and mortality in critically ill patients with acute neurological diseases: trauma and stroke differ from infection. Intensive Care Med 2015, 41(5):823–832.

5. Young PJ, Saxena M, Beasley R, Bellomo R, Bailey M, Pilcher D, Finfer S, Harrison D, Myburgh J, Rowan K: Early peak temperature and mortality in critically ill patients with or without infection. Intensive Care Med 2012.

6. Berman JD, Neva FA: Effect of temperature on multiplication of Leishmania amastigotes within human monocyte-derived macrophages in vitro. Am J Trop Med Hyg 1981, 30(2):318–321.

7. Mace TA, Zhong L, Kilpatrick C, Zynda E, Lee C-T, Capitano M, Minderman H, Repasky EA: Differentiation of CD8+ T cells into effector cells is enhanced by physiological range hyperthermia. Journal of Leukocyte Biology 2011, 90(5):951–962.

8. Chu CM, Tian SF, Ren GF, Zhang YM, Zhang LX, Liu GQ: Occurrence of temperature-sensitive influenza A viruses in nature. J Virol 1982, 41(2):353–359.

9. Moench LM: A Study of the Heat Sensitivity of the Meningoeoecus in Vitro within the Range of Therapeutic Temperatures. Journal of Laboratory and Clinical Medicine 1937, 22:665–676.

10. Small PM, Tauber MG, Hackbarth CJ, Sande MA: Influence of body temperature on bacterial growth rates in experimental pneumococcal meningitis in rabbits. Infection and immunity 1986, 52(2):484–487.

11. Mackowiak PA, Ruderman AE, Martin RM, Many WJ, Smith JW, Luby JP: Effects of physiologic variations in temperature on the rate of antibiotic-induced bacterial killing. American journal of clinical pathology 1981, 76(1):57–62.

12. Launey Y, Nesseler N, Mallédant Y, Seguin P: Clinical review: fever in septic ICU patients--friend or foe? Critical care (London, England) 2011, 15(3):222–222.

13. Doran TF, de Angelis C, Baumgardner RA, Mellits ED: Acetaminophen: more harm than good for chickenpox? J Pediatr 1989, 114(6):1045–1048.

14. Brandts CH, Ndjave M, Graninger W, Kremsner PG: Effect of paracetamol on parasite clearance time in Plasmodium falciparum malaria. Lancet 1997, 350(9079):704–709.

15. Stanley ED, Jackson GG, Panusarn C, Rubenis M, Dirda V: Increased virus shedding with aspirin treatment of rhinovirus infection. Jama 1975, 231(12):1248–1251.

16. Peters MJ, Woolfall K, Khan I, Deja E, Mouncey PR, Wulff J, Mason A, Agbeko RS, Draper ES, Fenn B et al: Permissive versus restrictive temperature thresholds in critically ill children with fever and infection: a multicentre randomized clinical pilot trial. Critical care (London, England) 2019, 23(1):69–69.

17. Evans SS, Repasky EA, Fisher DT: Fever and the thermal regulation of immunity: the immune system feels the heat. Nature reviews Immunology 2015, 15(6):335–349.

18. Lee CT, Zhong L, Mace TA, Repasky EA: Elevation in body temperature to fever range enhances and prolongs subsequent responsiveness of macrophages to endotoxin challenge. PLoS One 2012, 7(1):e30077.

19. Schulman CI, Namias N, Doherty J, Manning RJ, Li P, Elhaddad A, Lasko D, Amortegui J, Dy CJ, Dlugasch L et al: The effect of antipyretic therapy upon outcomes in critically ill patients: a randomized, prospective study. Surg Infect (Larchmt) 2005, 6(4):369–375.

20. Gozzoli V, Schottker P, Suter PM, Ricou B: Is it worth treating fever in intensive care unit patients? Preliminary results from a randomized trial of the effect of external cooling. Arch Intern Med 2001, 161 (1):121–123.

21. Young P, Saxena M, Bellomo R, Freebairn R, Hammond N, van Haren F, Holliday M, Henderson S, Mackle D, McArthur C et al: Acetaminophen for Fever in Critically Ill Patients with Suspected Infection. New England Journal of Medicine 2015, 373(23):2215–2224.

22. Zhang Z: Antipyretic therapy in critically ill patients with established sepsis: a trial sequential analysis. PLoS One 2015, 10(2):e0117279.

23. Dallimore J, Ebmeier S, Thayabaran D, Bellomo R, Bernard G, Schortgen F, Saxena M, Beasley R, Weatherall M, Young P: Effect of active temperature management on mortality in intensive care unit patients. Crit Care Resusc 2018, 20(2):150–163.

24. Drewry AM, Ablordeppey EA, Murray ET, Stoll CRT, Izadi SR, Dalton CM, Hardi AC, Fowler SA, Fuller BM, Colditz GA: Antipyretic Therapy in Critically Ill Septic Patients: A Systematic Review and Meta-Analysis. Critical care medicine 2017, 45(5):806–813.

25. Evans EM, Doctor RJ, Gage BF, Hotchkiss RS, Fuller BM, Drewry AM: The Association of Fever and Antipyretic Medication With Outcomes in Mechanically Ventilated Patients: A Cohort Study. Shock 2019, 52(2):152–159.

26. Raju TN: Hot brains: manipulating body heat to save the brain. Pediatrics 2006, 117(2):e320–321.

27. Epstein NN: Artificial Fever as a Therapeutic Procedure. Cal West Med 1936, 44(5):357–358.

28. Davis T: NICE guideline: feverish illness in children--assessment and initial management in children younger than 5 years. Archives of disease in childhood Education and practice edition 2013, 98(6):232–235.

29. van der Zee J: Heating the patient: a promising approach? Annals of Oncology 2002, 13(8):1173–1184.

30. Bull JMC: Clinical Practice of Whole-Body Hyperthermia: New Directions. In: Thermoradiotherapy and Thermochemotherapy. Medical Radiology (Diagnostic Imaging and Radiation Oncology). Berlin, Heidelberg: Springer; 1996.

31. Westermann AM, Grosen EA, Katschinski DM, Jäger D, Rietbroek R, Schink JC, Tiggelaar CL, Jäger E, Zum Vörde sive Vörding P, Neuman A et al: A pilot study of whole body hyperthermia and carboplatin in platinum-resistant ovarian cancer. European Journal of Cancer 2001, 37(9):1111–1117.

32. Robins HI, Dennis WH, Neville AJ, Shecterle LM, Martin PA, Grossman J, Davis TE, Neville SR, Gillis WK, Rusy BF: A nontoxic system for 41.8 degrees C whole-body hyperthermia: results of a Phase I study using a radiant heat device. Cancer research 1985, 45(8):3937–3944.

33. Shi H, Cao T, Connolly JE, Monnet L, Bennett L, Chapel S, Bagnis C, Mannoni P, Davoust J, Palucka AK et al: Hyperthermia Enhances CTL Cross-Priming. The Journal of Immunology 2006, 176(4):2134–2141.

34. Basu S, Srivastava PK: Fever-like temperature induces maturation of dendritic cells through induction of hsp90. International Immunology 2003, 15(9):1053–1061.

35. Tsan M-F, Gao B: Heat shock proteins and immune system. Journal of Leukocyte Biology 2009, 85(6):905–910.

36. Foxman EF, Storer JA, Fitzgerald ME, Wasik BR, Hou L, Zhao H, Turner PE, Pyle AM, Iwasaki A: Temperature-dependent innate defense against the common cold virus limits viral replication at warm temperature in mouse airway cells. Proceedings of the National Academy of Sciences 2015, 112(3):827–832.

37. Ping CK: Rapid response to: Graphic Outbreak of severe acute respiratory syndrome in Hong Kong Special Administrative Region: case report. BMJ 2003, 326(850).

38. Laporte M, Stevaert A, Raeymaekers V, Boogaerts T, Nehlmeier I, Chiu W, Benkheil M, Vanaudenaerde B, Pöhlmann S, Naesens L: Hemagglutinin Cleavability, Acid Stability, and Temperature Dependence Optimize Influenza B Virus for Replication in Human Airways. Journal of Virology 2019, 94(1):e01430–01419.

39. Zou L, Ruan F, Huang M, Liang L, Huang H, Hong Z, Yu J, Kang M, Song Y, Xia J et al: SARS-CoV-2 Viral Load in Upper Respiratory Specimens of Infected Patients. N Engl J Med 2020, 382(12):1177–1179.

40. Chan KH, Peiris JS, Lam SY, Poon LL, Yuen KY, Seto WH: The Effects of Temperature and Relative Humidity on the Viability of the SARS Coronavirus. Advances in virology 2011, 2011:734690.

41. Wang W, Xu Y, Gao R, Lu R, Han K, Wu G, Tan W: Detection of SARS-CoV-2 in Different Types of Clinical Specimens. Jama 2020.

42. He J, Tao H, Yan Y, Huang S-Y, Xiao Y: Molecular mechanism of evolution and human infection with the novel coronavirus (2019-nCoV). bioRxiv 2020:2020.2002.2017.952903.

43. Zhou F, Yu T, Du R, Fan G, Liu Y, Liu Z, Xiang J, Wang Y, Song B, Gu X et al: Clinical course and risk factors for mortality of adult inpatients with COVID-19 in Wuhan, China: a retrospective cohort study. The Lancet 2020, 395(10229):1054–1062.

44. Goury A, Poirson F, Chaput U, Voicu S, Garcon P, Beeken T, Malissin I, Kerdjana L, Chelly J, Vodovar D et al: Targeted Temperature Management Using The "Esophageal Cooling Device" After Cardiac Arrest (The COOL Study): A feasibility and safety study. Resuscitation 2017, 121:54–61.

45. Hegazy AF, Lapierre DM, Butler R, Martin J, Althenayan E: The esophageal cooling device: A new temperature control tool in the intensivist’s arsenal. Heart & Lung: The Journal of Acute and Critical Care 2017, 46(3):143–148.

46. Markota A, Fluher J, Kit B, Balazic P, Sinkovic A: The introduction of an esophageal heat transfer device into a therapeutic hypothermia protocol: A prospective evaluation. Am J Emerg Med 2016, 34(4):741–745.

47. Khan I, Haymore J, Barnaba B, Armahizer M, Melinosky C, Bautista MA, Blaber B, Chang WT, Parikh G, Motta M et al: Esophageal Cooling Device Versus Other Temperature Modulation Devices for Therapeutic Normothermia in Subarachnoid and Intracranial Hemorrhage. Ther Hypothermia Temp Manag 2018, 8(1):53–58.

48. Williams D, Leslie G, Kyriazis D, O’Donovan B, Bowes J, Dingley J: Use of an Esophageal Heat Exchanger to Maintain Core Temperature during Burn Excisions and to Attenuate Pyrexia on the Burns Intensive Care Unit. Case Reports in Anesthesiology 2016, 2016:6.

49. Kalasbail P, Makarova N, Garrett F, Sessler DI: Heating and Cooling Rates With an Esophageal Heat Exchange System. Anesth Analg 2018, 126(4):1190–1195.

50. Bhatti F, Naiman M, Tsarev A, Kulstad E: Esophageal Temperature Management in Patients Suffering from Traumatic Brain Injury. Ther Hypothermia Temp Manag 2019.

51. Martin KR, Naiman M, Espinoza M: Using Esophageal Temperature Management to Treat Severe Heat Stroke: A Case Report. J Neurosci Nurs 2019.

52. Hegazy AF, Lapierre DM, Butler R, Althenayan E: Temperature control in critically ill patients with a novel esophageal cooling device: a case series. BMC anesthesiology 2015, 15:152.

53. Markota A, Košir AS, Balažič P, Živko I, Sinkovič A: A Novel Esophageal Heat Transfer Device for Temperature Management in an Adult Patient with Severe Meningitis. Journal of Emergency Medicine 2017, 52(1):e27–e28

